# Determinants of mpox vaccine acceptance among peri-urban residents in Sekondi-Takoradi, Western Region of Ghana: A community-based cross-sectional study

**DOI:** 10.64898/2026.11.10.26367771

**Authors:** Gifty Amugi, Emmanuel Mensah Baah, Herbert Yankey, Isaac Kwarkye, Pius Mensah, Kofi Jean Claude, Dennis Jubin, Samuel Kaba Akoriyea, Franklin Asiedu Bekoe, Dennis Laryea, Azumah Abdul-Tawab, Joshua Arthur, Selorm Kutsoati, Joseph Oliver-Commey, Naziru Tanko Mohammed

## Abstract

Mpox re-emerged as a global public health concern in 2022, and the Western Region of Ghana became the epicentre of the country’s largest recorded outbreak in 2025. Ahead of a sub-national vaccination campaign launched in October 2025, no study had examined the determinants of mpox vaccine acceptance among peri-urban residents of the region using a theoretically grounded framework. This study determined the sociodemographic and Health Belief Model-based determinants of mpox vaccine acceptance among peri-urban residents of the Sekondi-Takoradi Metropolis, Western Region, Ghana. A community-based cross-sectional study was conducted among 580 adults in six purposively selected peri-urban areas and their surrounding communities between July and August 2025, before the vaccination campaign began. Vaccine acceptance was measured as a three-category outcome (accept, hesitant, refuse) and analysed using chi-square tests and multinomial logistic regression adjusted for sociodemographic characteristics, Health Belief Model constructs, and peri-urban area. Overall, 75.7% of respondents were willing to accept the vaccine, 12.9% were hesitant and 11.4% would refuse. Perceived vaccine safety was the strongest determinant: respondents who considered the vaccine very safe had markedly lower adjusted odds of hesitancy (adjusted odds ratio 0.02, 95% confidence interval 0.00–0.12) and refusal (0.03, 0.01–0.13) than those who considered it unsafe. Uncertainty about vaccine effectiveness was associated with hesitancy (9.83, 1.95–49.50), and belief in its effectiveness with lower odds of refusal (0.22, 0.06–0.86). Higher educational attainment was associated with lower odds of hesitancy, being a student or employed with lower odds of refusal, and acceptance differed between peri-urban areas (55.1% to 95.6%). Knowledge of mpox transmission was associated with acceptance in bivariate analysis (p < 0.001) but not after adjustment. Risk communication that addresses vaccine safety and effectiveness, rather than generic disease-awareness messaging alone, is likely to be most effective in improving mpox vaccine acceptance in this population, with unemployed and less-educated residents warranting targeting. As a pre-campaign baseline collected during an active outbreak, these findings can inform the vaccination response in the Western Region and preparedness in comparable peri-urban West African settings.

## Introduction

### Background

Mpox (formerly monkeypox) is a zoonotic disease caused by the monkeypox virus, a member of the Orthopoxvirus genus within the Poxviridae family, first identified in 1958 during an outbreak among laboratory monkeys, with the first human case documented in 1970 in the Democratic Republic of the Congo [1]. The virus is classified into two clades: Clade I, historically endemic to Central Africa and associated with greater disease severity, and Clade II, endemic to West Africa and generally associated with milder illness [1,2]. Although mpox had circulated at low levels in endemic African countries for decades, the disease gained unprecedented global attention in May 2022, when Clade IIb strains began spreading across multiple non-endemic countries, prompting the World Health Organization (WHO) to declare a Public Health Emergency of International Concern in July 2022 [2,3]. A second Public Health Emergency of International Concern was declared in August 2024 following the emergence and spread of the more transmissible Clade Ib strain in the Democratic Republic of the Congo and neighbouring countries [1]. By late 2025, the WHO had recorded tens of thousands of confirmed cases across the African region alone, with the Democratic Republic of Congo, Liberia, Ghana, Kenya, and Uganda reporting the highest recent case burdens [4].

Ghana has not been insulated from this resurgence. Following sporadic cases in late 2024 and early 2025, a substantial outbreak began in May 2025, with the Western Region rapidly emerging as the country’s epicentre, at one point accounting for over 70% of national confirmed cases [5]. Within the Western Region, the Sekondi-Takoradi Metropolis recorded a sustained rise in suspected and confirmed cases through the third quarter of 2025, prompting the Ghana Health Service, in partnership with the WHO and the Ministry of Health, to launch a sub-national vaccination campaign targeting the most affected districts from October 2025 [5,6]. Vaccination remains one of the most effective tools for interrupting mpox transmission and protecting at-risk populations [7], yet the success of any vaccination campaign depends fundamentally on population-level willingness to be vaccinated [8].

### Vaccine hesitancy and acceptance as a global public health challenge

Vaccine hesitancy, defined as the delay in acceptance or refusal of vaccination despite the availability of vaccination services [9], has been identified as a persistent obstacle to mpox outbreak control worldwide, though its magnitude varies considerably by population and context. Among healthcare workers, studies have reported acceptance ranging from as low as 8.8% in the Czech Republic [10] to over 90% in China [11], while among men who have sex with men, a population prioritised for vaccination in several countries, acceptance has generally been higher, ranging from 81.5% in the Netherlands [12] to 86.2% in China [13]. Among general population samples, findings have been similarly heterogeneous: moderate acceptance has been reported in the Middle East [14] and Peru [15], while acceptance in Romania has remained persistently low despite a modest upward trend between 2022 and 2025 [16]. This variability underscores that mpox vaccine acceptance is not a fixed or universal quantity but is instead shaped by context-specific social, cultural, and epidemiological factors that must be understood locally rather than assumed from findings elsewhere.

### Mpox vaccine acceptance in Ghana and West Africa

Empirical evidence on mpox vaccine acceptance within Ghana specifically remains limited and, where available, points to considerable hesitancy. Ghazy et al. [17], surveying a national online sample of Ghanaians in late 2022, found that only 46.1% intended to accept the mpox vaccine, with urban residence, prior refusal of the COVID-19 vaccine, and low vaccine confidence identified as key determinants. Using more recent cross-sectional data collected across four major Ghanaian cities, Braimah et al. [18] reported an even lower acceptance rate of approximately 32%, with prior vaccination history emerging as a critical predictor. Comparable studies from neighbouring Nigeria have similarly documented moderate hesitancy: Ghazy et al. [19] found low vaccine confidence and high perceived constraints among Nigerian healthcare workers, while Iliyasu et al. [20], in a mixed-methods study among the general public in Kano, found that only half of respondents were willing to accept the vaccine despite near-universal awareness of the disease.

This body of evidence, while informative, shares three important limitations relative to the present context. First, existing Ghanaian studies were conducted online or in urban centres, predominantly using convenience or snowball sampling, and none has focused specifically on peri-urban populations. Second, all were conducted either before the 2025 outbreak or during its earliest phase, when mpox in Ghana remained a comparatively abstract, low-visibility threat rather than a lived, community-level emergency. Third, each measured vaccine acceptance as a binary outcome, collapsing hesitancy and outright refusal into a single category despite growing recognition that these represent conceptually distinct psychological states [18]. A broader literature on COVID-19 vaccine acceptance in Ghana, including work by Aram et al. [22], Tabong et al. [23], Aberese-Ako et al. [24], and Aninagyei et al. [25], the latter conducted in the Western Region itself, offers useful methodological precedent but cannot substitute for disease-and context-specific evidence, given that determinants of vaccine acceptance are known to vary meaningfully across different vaccines and outbreak contexts [21].

### Theoretical framework: The Health Belief Model

The Health Belief Model (HBM) is among the most widely applied theoretical frameworks in vaccine acceptance research, positing that health-protective behaviour is shaped by an individual’s perceived susceptibility to a health threat, perceived severity of that threat, perceived benefits of the recommended action, perceived barriers to taking that action, and exposure to cues that prompt action, with self-efficacy incorporated in later extensions of the model [26]. The HBM has been applied directly to mpox vaccine acceptance in several international settings. Riad et al. [10] found that cues to action and perceived susceptibility were the strongest predictors of mpox vaccine acceptance among Czech healthcare workers, while Mohamed et al. [27], applying the full six-construct model among healthcare workers across three Gulf countries, identified perceived benefits and self-efficacy as the dominant predictors. Among Belarusian healthcare professionals, Riad et al. [28] similarly found that perceived benefits and cues to action most strongly predicted willingness to vaccinate.

Although the HBM has not previously been applied to mpox vaccine acceptance in Ghana, it has been validated for other health behaviours within the Ghanaian context, lending support to its theoretical appropriateness for this population. Tweneboah-Koduah [29] found that HBM constructs explained nearly 69% of the variance in breast cancer protective behaviours among Ghanaian women, Kyei-Gyamfi et al. [30] applied the model to condom use among internal migrant girls in Ghana, and Bawuah et al. [31] drew on HBM-informed prior work to explain cervical cancer screening uptake among Ghanaian women. This establishes a clear and consequential gap: no known study has yet applied the Health Belief Model to mpox vaccine acceptance within Ghana, despite the model’s demonstrated relevance both to mpox vaccination behaviour internationally and to other health behaviours domestically.

### Residence and vaccine acceptance

Place of residence has been repeatedly identified as a determinant of vaccine acceptance, though the direction of this association is inconsistent across settings. Some studies report higher hesitancy in urban areas: Cissé et al. [35] found significantly higher COVID-19 vaccine hesitancy in urban compared to rural areas of Burkina Faso, a pattern also observed across three successive waves of a national Chinese survey [32]. Others report the opposite pattern: in the United States, rural residents showed lower HBM-measured risk perception and lower intention to accept the mpox vaccine than urban residents [33], and rural adolescents showed significantly lower HPV vaccination uptake than their urban counterparts [34]. This inconsistency suggests that residence-based effects are context-dependent rather than universal and reinforces the recommendation of Oladapo et al. [36] that vaccine promotion strategies in low-and middle-income countries be deliberately tailored to peri-urban and semi-urban communities rather than assumed to mirror either fully urban or fully rural patterns. Peri-urban populations, which share some characteristics of both urban and rural settings while remaining distinct from each, thus represent a specific and, in the Ghanaian mpox context, entirely unstudied population.

### Rationale and objectives

Despite the Western Region of Ghana emerging as the epicentre of the country’s largest recorded mpox outbreak, and with a sub-national vaccination campaign about to be launched in the Sekondi-Takoradi Metropolis, no empirical study has examined the determinants of mpox vaccine acceptance among residents of this specific, heavily affected area. Existing Ghanaian evidence on mpox vaccine acceptance is drawn from national or urban online samples collected before or during the earliest phase of the outbreak, measures acceptance as a binary outcome that cannot distinguish hesitancy from outright refusal, and has not applied a theoretical framework capable of explaining why acceptance varies as it does. This leaves public health authorities in the Western Region without a locally grounded, theoretically informed evidence base to guide the forthcoming vaccination response, a gap this study addresses directly.

This study offers three distinct contributions. First, by collecting data during an active, geographically concentrated outbreak, it provides a rare within-outbreak account of vaccine-related beliefs under conditions of real rather than hypothetical threat, addressing a temporal limitation of the existing Ghanaian literature. Second, by focusing specifically on peri-urban communities within the Sekondi-Takoradi Metropolis, it addresses a population that existing residence-based vaccine acceptance research, in Ghana and elsewhere, has not directly examined. Third, by applying the Health Belief Model and modeling acceptance as a three-category outcome distinguishing acceptance, hesitancy, and refusal, it offers a more theoretically grounded and analytically precise account of vaccine decision-making than prior binary-outcome studies in this context have provided. The findings are intended to directly inform risk communication and vaccination strategy in the Western Region response, and to contribute reusable, context-specific evidence for future mpox outbreak preparedness in similar peri-urban West African settings.

The general objective of this study was to determine the Health Belief Model-based and sociodemographic determinants of mpox vaccine acceptance among peri-urban residents of the Sekondi-Takoradi Metropolis, Western Region of Ghana. The specific objectives were to (i) assess the level of knowledge of mpox and its transmission among peri-urban residents of the metropolis; (ii) determine the level of mpox vaccine acceptance; (iii) assess the Health Belief Model constructs (perceived susceptibility, perceived severity, perceived benefits, perceived barriers, and cues to action) regarding mpox vaccination; and (iv) determine the sociodemographic and Health Belief Model determinants independently associated with mpox vaccine acceptance.

## Materials and methods

### Study design and setting

This was a community-based, cross-sectional study conducted among adult residents of peri-urban communities within the Sekondi-Takoradi Metropolis (STM), Western Region, Ghana, between July and August 2025. Data collection took place during an active regional mpox outbreak and before the sub-national mpox vaccination campaign was launched in October 2025 [6]. The study examined mpox-related knowledge, Health Belief Model constructs, and stated willingness to accept the mpox vaccine; it did not assess actual vaccination uptake.

### Study population and eligibility

The study population comprised adults aged 18 years and above who had lived permanently in a peri-urban community of the STM for more than one year. Peri-urban communities were identified using the Ghana Statistical Service classification. Visitors and non-residents, people who were too ill or otherwise unable to give informed consent, and individuals younger than 18 years were excluded.

### Sample size determination

The minimum required sample size was estimated using the Cochran formula for proportions: n = Z²p(1−p)/d², where Z = 1.96 (95% confidence level), p = 0.50 (a conservative estimate, in the absence of prior community-based mpox vaccine acceptance data in Ghana), and d = 0.05, yielding a minimum of 385 participants. Allowing for a 10% non-response rate, the adjusted target was 424 participants. In total, 580 eligible adults were interviewed, and the response rate was 98%.

### Sampling technique

A two-stage sampling approach was used. In the first stage, six peri-urban areas of the STM, all classified as peri-urban by the Ghana Statistical Service, were purposively selected: Kojokrom, Essikado, Adiembra, Diabene, Essipong, and Kansaworodo. The six areas were selected on the basis of the degree and pace of urban expansion within them and the size of their resident populations, so that, of all the peri-urban areas in the STM, those included were the ones where a vaccination campaign would reach the largest number of eligible people [37].

In the second stage, trained data collectors visited each selected area and the communities immediately surrounding it, and approached adults in their homes and in the immediate vicinity; eligible adults who were available and consented were interviewed. Because respondents at this stage were recruited on the basis of availability rather than random selection, this stage constituted convenience sampling. Each respondent was assigned to a peri-urban area using the GPS coordinates recorded in KoboCollect at the time of interview. The final sample comprised 183 respondents from Kojokrom, 127 from Essikado, 107 from Adiembra, 87 from Diabene, 46 from Essipong, and 30 from Kansaworodo (total n = 580; response rate 98%).

### Theoretical framework

This study was guided by the Health Belief Model (HBM), operationalised across five constructs: perceived susceptibility, perceived severity, perceived benefits, perceived barriers, and cues to action. Self-efficacy, while part of the extended HBM, was not directly measured by this instrument and is not included as an operational construct in this study.

### Data collection instrument

Data were collected using a structured, interviewer-administered questionnaire programmed in KoboCollect and completed electronically. The questionnaire comprised six sections: (1) sociodemographic characteristics; (2) knowledge of mpox and its transmission; (3) awareness of and exposure to information about the mpox vaccine; (4) HBM construct items; (5) vaccine acceptance and perceived barriers; and (6) factors that would increase willingness to vaccinate. Questions on transmission and on the vaccine were asked only of respondents who had heard of mpox. The questionnaire was pre-tested among 30 respondents in 10 communities outside the study area, and no changes were required before data collection. Data were collected by 24 trained data collectors, comprising community health volunteers, community health nurses, health promotion officers, and other public health officers, who were trained in the study objectives, the questionnaire, and informed consent procedures before fieldwork. Interviews were conducted in English, Fante, or Twi, according to the language each respondent understood best; data collectors fluent in these languages translated the questions orally, using standardised wording agreed during training.

### Variable measurement

#### Outcome variable

Willingness to accept the mpox vaccine was measured with a single item and treated as a three-level categorical outcome: accept, hesitant (“not sure”), and refuse, consistent with the definition of the WHO Strategic Advisory Group of Experts Working Group on Vaccine Hesitancy, which treats hesitancy as encompassing delay or uncertainty as distinct from outright refusal.

#### Knowledge score

Among respondents who had heard of mpox, a composite transmission-knowledge score (range 0–4) was constructed by summing correctly identified transmission routes (skin-to-skin contact, sexual contact, contact with contaminated objects, consumption of bush meat); selection of the distractor item (“mosquito bite”) was recorded separately as a misconception indicator. Scores were categorised as poor (0–1), moderate (2–3), or good (4). Respondents who had not heard of mpox (n = 85) were not asked these questions and were analysed as a separate category.

#### HBM constructs

Perceived susceptibility (whether respondents considered themselves at risk of contracting mpox), perceived severity (concern about becoming infected), and perceived benefits (belief that the vaccine can prevent the disease) were each measured with a single categorical item. Perceived barriers were measured with a single item on perceived vaccine safety (very safe, somewhat safe, don’t know, not safe) and a count of endorsed concerns (side effects, lack of information, mistrust in vaccines, belief that mpox is not serious, and religious or cultural reasons; range 0–5). Cues to action were operationalised as a count of endorsed factors that would increase willingness to vaccinate (more information from health authorities, seeing others vaccinated, assurance of safety and effectiveness, and community leader endorsement; range 0–4).

#### Peri-urban area

Each respondent’s peri-urban area (Kojokrom, Essikado, Adiembra, Diabene, Essipong, or Kansaworodo) was recorded as a six-category variable.

## Statistical analysis

Data were analysed in Python (version 3.13) using pandas (version 3.0), SciPy (version 1.18), and statsmodels (version 0.15); the analysis code is provided in S1 Code. Descriptive statistics are presented as frequencies and percentages for categorical variables and means with standard deviations for continuous variables. Bivariate associations between candidate determinants and vaccine acceptance status were assessed using the chi-square test of independence; where more than 20% of expected cell frequencies were below 5, a Monte Carlo permutation test (10,000 permutations) was used in place of the asymptotic p-value. Multivariable analysis used multinomial logistic regression with acceptance as the reference outcome category. Predictors were specified a priori from the HBM and sociodemographic characteristics: sex, age (continuous, in years), education, occupation (student, employed, unemployed/retired), marital status (single, ever married), religion (Christian, non-Christian), perceived susceptibility, perceived severity, perceived benefits, perceived vaccine safety, barrier count, and cue count. Peri-urban area was included as a fixed effect to account for differences between areas and the clustering of respondents within them. Crude odds ratios (OR) were estimated from models containing each predictor alone, and adjusted odds ratios (aOR) with 95% confidence intervals (CI) from the full model. Model fit was assessed with McFadden’s pseudo-R² and the likelihood ratio test, and the contributions of perceived vaccine safety, peri-urban area, and transmission knowledge were assessed with likelihood ratio tests comparing nested models.

Multicollinearity was assessed with variance inflation factors. There were no missing data for variables in the regression model; transmission-knowledge items were skipped by design for respondents who had not heard of mpox. Statistical significance was set at p < 0.05.

## Ethics statement

This study was conducted in accordance with the principles of the Declaration of Helsinki. Ethical approval was obtained from the University Research Ethics Committee of Takoradi Technical University, Takoradi, Ghana (reference number TTU/UREC/26/097, 17 June 2025), and permission was obtained from the Sekondi-Takoradi Metropolitan Health Directorate. Informed consent was obtained verbally: the consent statement was read aloud to each respondent in the language of the interview, and the respondent’s decision to participate or decline was recorded electronically in KoboCollect before any questions were asked. Verbal consent was used because data were collected electronically in an anonymous, minimal-risk survey that included respondents with limited literacy, and this procedure was approved by the ethics committee. No personal identifiers were collected.

## Results

### Sociodemographic characteristics of respondents

A total of 580 peri-urban residents of the Sekondi-Takoradi Metropolis completed the survey. The majority of respondents were female (406, 70.0%), with a mean age of 33.5 years (SD 12.6; range 18–75). Most respondents had attained secondary education (241, 41.6%) or tertiary education (205, 35.3%), while 134 (23.1%) had no formal education, primary education only, or Junior High School education. Students constituted the largest occupational group (192, 33.1%), followed by the self-employed (149, 25.7%) and government/public sector workers (143, 24.7%). The majority were single (343, 59.1%), and most identified as Christian (511, 88.1%), with 61 (10.5%) identifying as Muslim. Respondents were drawn from Kojokrom (183, 31.6%), Essikado (127, 21.9%), Adiembra (107, 18.4%), Diabene (87, 15.0%), Essipong (46, 7.9%), and Kansaworodo (30, 5.2%) (Table 1).

**Table 1.**
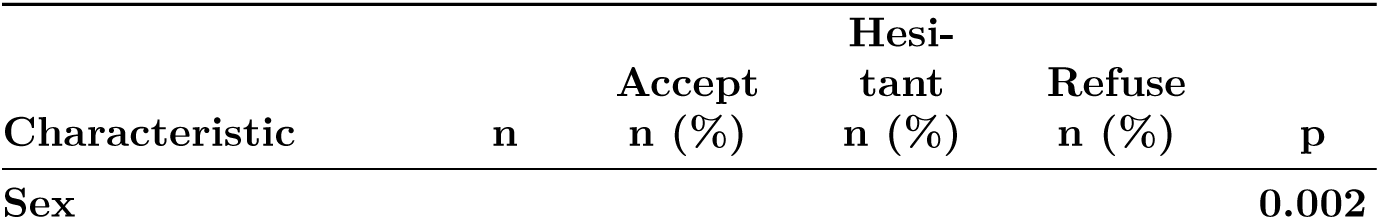
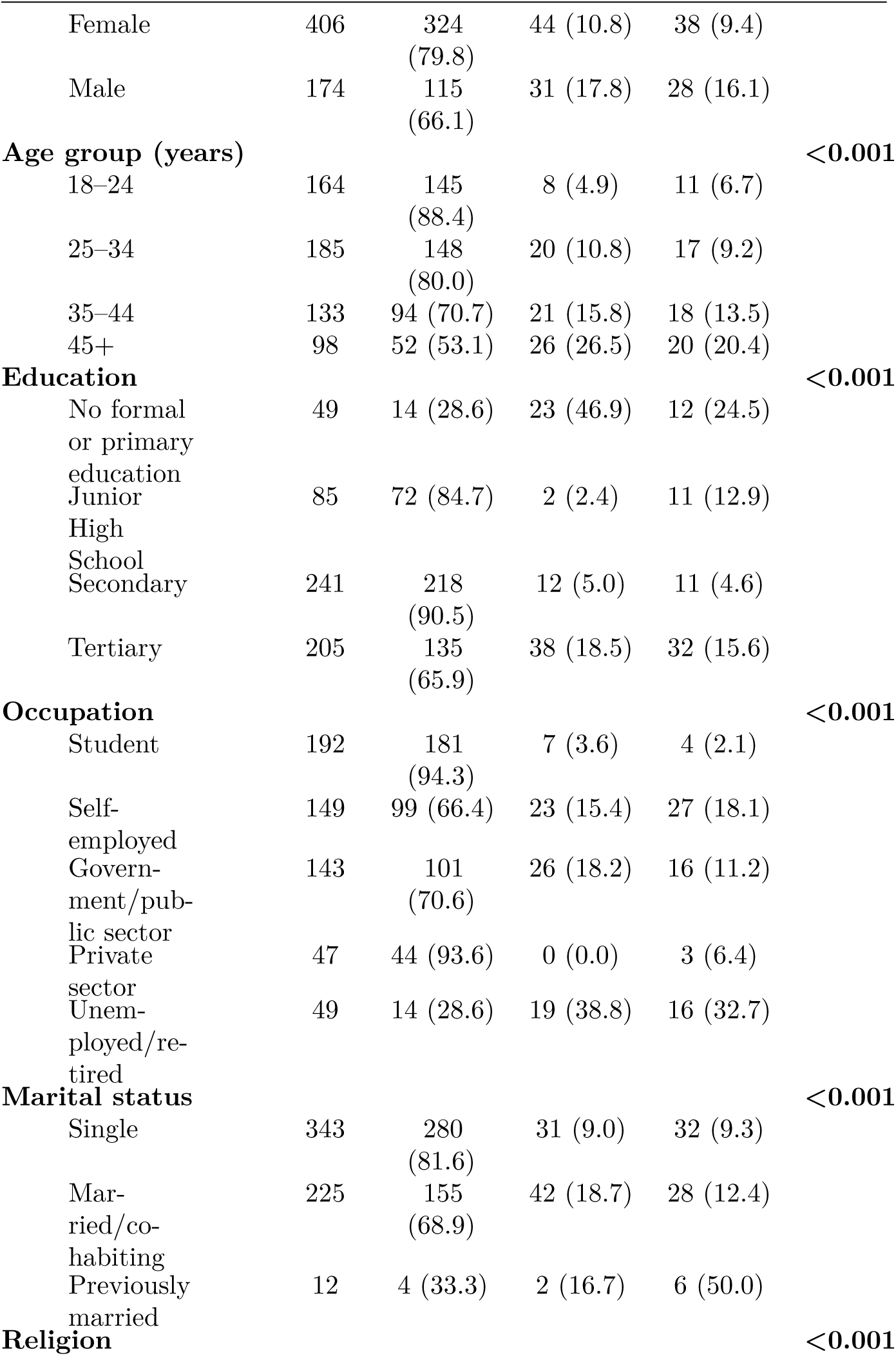
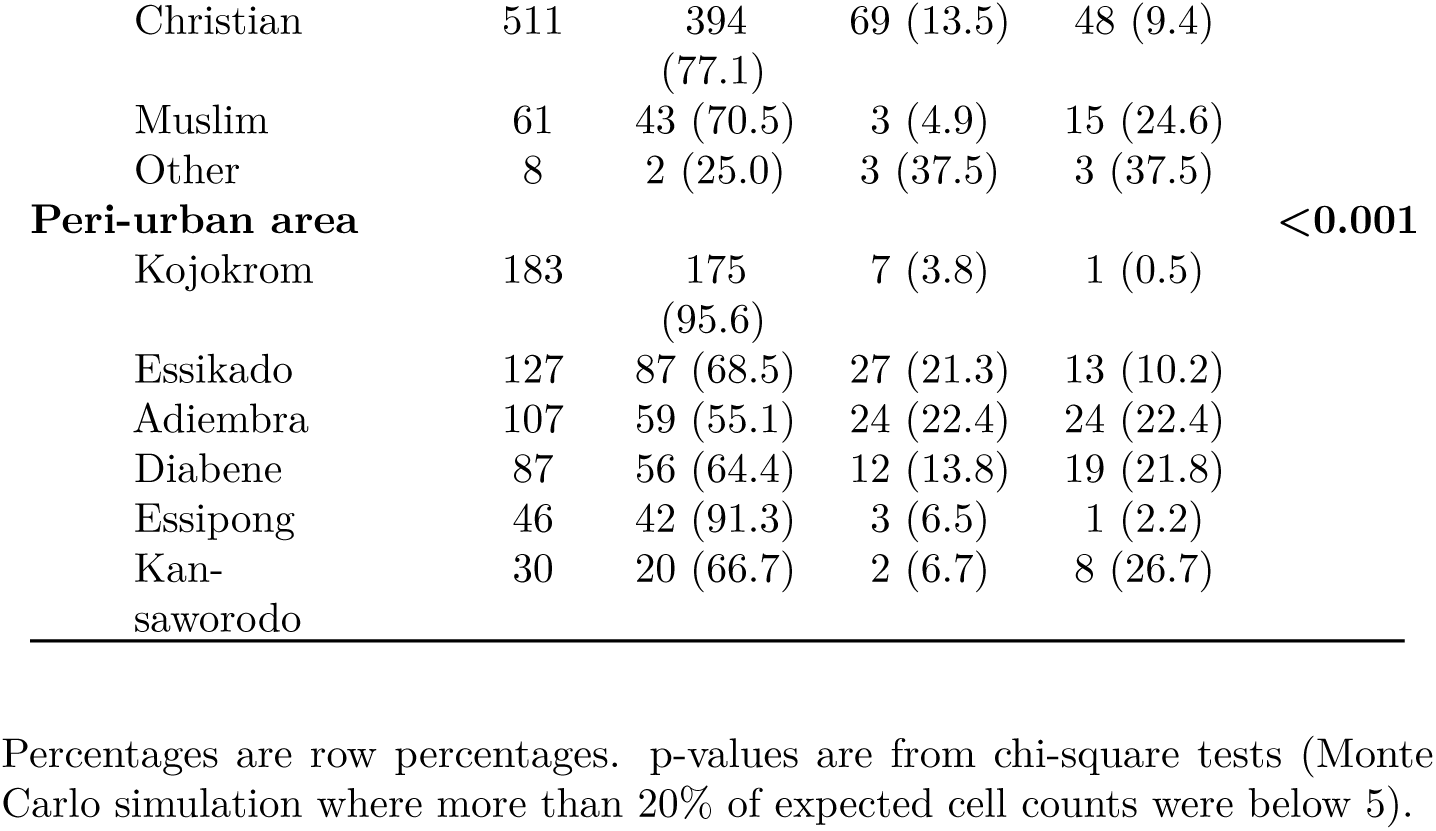
Sociodemographic characteristics of respondents by mpox vaccine acceptance status (n = 580).

| **Characteristic** | **n** | **Accept n (%)** | **Hesi-tant n (%)** | **Refuse n (%)** | **p** |
|---|---|---|---|---|---|
| **Sex** |  |  |  |  | **0.002** |
| Female | 406 | 324 (79.8) | 44 (10.8) | 38 (9.4) |  |
| Male | 174 | 115 (66.1) | 31 (17.8) | 28 (16.1) |  |
| **Age group (years)** |  |  |  |  | **<0.001** |
| 18–24 | 164 | 145 (88.4) | 8 (4.9) | 11 (6.7) |  |
| 25–34 | 185 | 148 (80.0) | 20 (10.8) | 17 (9.2) |  |
| 35–44 | 133 | 94 (70.7) | 21 (15.8) | 18 (13.5) |  |
| 45+ | 98 | 52 (53.1) | 26 (26.5) | 20 (20.4) |  |
| **Education** |  |  |  |  | **<0.001** |
| No formal or primary education | 49 | 14 (28.6) | 23 (46.9) | 12 (24.5) |  |
| Junior High School | 85 | 72 (84.7) | 2 (2.4) | 11 (12.9) |  |
| Secondary | 241 | 218 (90.5) | 12 (5.0) | 11 (4.6) |  |
| Tertiary | 205 | 135 (65.9) | 38 (18.5) | 32 (15.6) |  |
| **Occupation** |  |  |  |  | **<0.001** |
| Student | 192 | 181 (94.3) | 7 (3.6) | 4 (2.1) |  |
| Self-employed | 149 | 99 (66.4) | 23 (15.4) | 27 (18.1) |  |
| Govern-ment/pub-lic sector | 143 | 101 (70.6) | 26 (18.2) | 16 (11.2) |  |
| Private sector | 47 | 44 (93.6) | 0 (0.0) | 3 (6.4) |  |
| Unem-ployed/re-tired | 49 | 14 (28.6) | 19 (38.8) | 16 (32.7) |  |
| **Marital status** |  |  |  |  | **<0.001** |
| Single | 343 | 280 (81.6) | 31 (9.0) | 32 (9.3) |  |
| Mar-ried/co-habiting | 225 | 155 (68.9) | 42 (18.7) | 28 (12.4) |  |
| Previously married | 12 | 4 (33.3) | 2 (16.7) | 6 (50.0) |  |
| **Religion** |  |  |  |  | **<0.001** |
| Christian | 511 | 394 (77.1) | 69 (13.5) | 48 (9.4) |  |
| Muslim | 61 | 43 (70.5) | 3 (4.9) | 15 (24.6) |  |
| Other | 8 | 2 (25.0) | 3 (37.5) | 3 (37.5) |  |
| **Peri-urban area** |  |  |  |  | **<0.001** |
| Kojokrom | 183 | 175 (95.6) | 7 (3.8) | 1 (0.5) |  |
| Essikado | 127 | 87 (68.5) | 27 (21.3) | 13 (10.2) |  |
| Adiembra | 107 | 59 (55.1) | 24 (22.4) | 24 (22.4) |  |
| Diabene | 87 | 56 (64.4) | 12 (13.8) | 19 (21.8) |  |
| Essipong | 46 | 42 (91.3) | 3 (6.5) | 1 (2.2) |  |
| Kan-saworodo | 30 | 20 (66.7) | 2 (6.7) | 8 (26.7) |  |

Percentages are row percentages. p-values are from chi-square tests (Monte Carlo simulation where more than 20% of expected cell counts were below 5).

### Knowledge and awareness of mpox and the mpox vaccine

Most respondents (495, 85.3%) reported having heard of mpox prior to the survey; the 85 who had not were not asked about transmission routes. Among the 495 aware respondents, the mean transmission-knowledge score was 2.8 out of 4 (SD 1.3), with 250 (50.5%) classified as having good knowledge, 126 (25.5%) moderate, and 119 (24.0%) poor. Notably, 141 aware respondents (28.5%) endorsed mosquito bites as a transmission route, a documented misconception, indicating that gaps in accurate knowledge persisted even among a broadly aware population. Of the 495 respondents aware of mpox, 349 (70.5%) knew a vaccine existed, 112 (22.6%) did not, and 34 (6.9%) were unsure.

### Health Belief Model construct distributions

Across the sample, 429 respondents (74.0%) considered themselves at risk of contracting mpox (perceived susceptibility), and 454 (78.3%) reported being very concerned about infection (perceived severity). Regarding perceived benefits, 449 (77.4%) believed the vaccine could effectively prevent the disease, while 94 (16.2%) were unsure and 37 (6.4%) did not believe it was effective. For perceived barriers, belief in vaccine safety was generally high (381, 65.7% considered it “very safe”), though 34 (5.9%) considered it unsafe. The most frequently endorsed specific barriers were fear of side effects (459, 79.1%), mistrust in vaccines (367, 63.3%), and lack of information (330, 56.9%); religious or cultural objections were comparatively rare (65, 11.2%). On cues to action, more information from health authorities (459, 79.1%) and assurance of safety and effectiveness (436, 75.2%) were the factors respondents most frequently indicated would increase their willingness to vaccinate (Table 2).

**Table 2.**
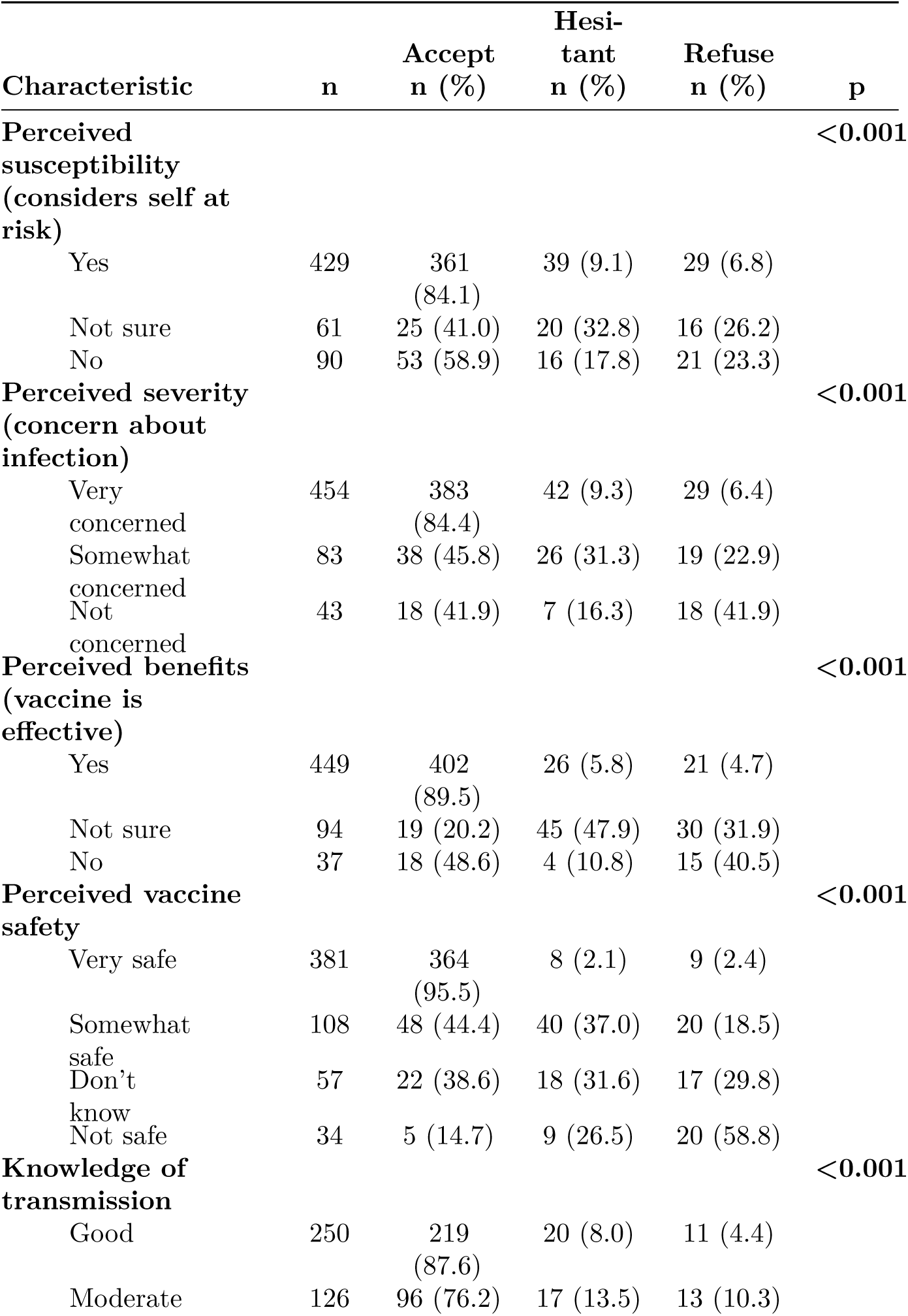
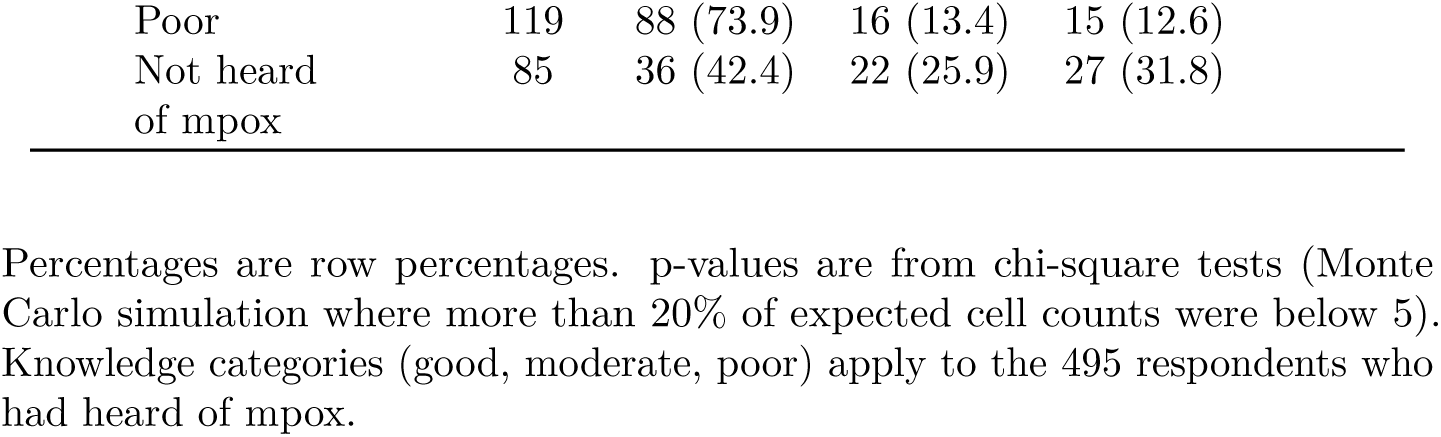
Health Belief Model constructs and knowledge of transmission by mpox vaccine acceptance status (n = 580).

| **Characteristic** | **n** | **Accept n (%)** | **Hesi-tant n (%)** | **Refuse n (%)** | **p** |
|---|---|---|---|---|---|
| **Perceived susceptibility (considers self at risk)** |  |  |  |  | **<0.001** |
| Yes | 429 | 361 (84.1) | 39 (9.1) | 29 (6.8) |  |
| Not sure | 61 | 25 (41.0) | 20 (32.8) | 16 (26.2) |  |
| No | 90 | 53 (58.9) | 16 (17.8) | 21 (23.3) |  |
| **Perceived severity (concern about infection)** |  |  |  |  | **<0.001** |
| Very concerned | 454 | 383 (84.4) | 42 (9.3) | 29 (6.4) |  |
| Somewhat concerned | 83 | 38 (45.8) | 26 (31.3) | 19 (22.9) |  |
| Not concerned | 43 | 18 (41.9) | 7 (16.3) | 18 (41.9) |  |
| **Perceived benefits (vaccine is effective)** |  |  |  |  | **<0.001** |
| Yes | 449 | 402 (89.5) | 26 (5.8) | 21 (4.7) |  |
| Not sure | 94 | 19 (20.2) | 45 (47.9) | 30 (31.9) |  |
| No | 37 | 18 (48.6) | 4 (10.8) | 15 (40.5) |  |
| **Perceived vaccine safety** |  |  |  |  | **<0.001** |
| Very safe | 381 | 364 (95.5) | 8 (2.1) | 9 (2.4) |  |
| Somewhat safe | 108 | 48 (44.4) | 40 (37.0) | 20 (18.5) |  |
| Don't know | 57 | 22 (38.6) | 18 (31.6) | 17 (29.8) |  |
| Not safe | 34 | 5 (14.7) | 9 (26.5) | 20 (58.8) |  |
| **Knowledge of transmission** |  |  |  |  | **<0.001** |
| Good | 250 | 219 (87.6) | 20 (8.0) | 11 (4.4) |  |
| Moderate | 126 | 96 (76.2) | 17 (13.5) | 13 (10.3) |  |
| Poor | 119 | 88 (73.9) | 16 (13.4) | 15 (12.6) |  |
| Not heard of mpox | 85 | 36 (42.4) | 22 (25.9) | 27 (31.8) |  |

Percentages are row percentages. p-values are from chi-square tests (Monte Carlo simulation where more than 20% of expected cell counts were below 5). Knowledge categories (good, moderate, poor) apply to the 495 respondents who had heard of mpox.

### Vaccine acceptance status

Overall, 439 respondents (75.7%) indicated they would accept the mpox vaccine if offered, 75 (12.9%) were hesitant (“not sure”), and 66 (11.4%) indicated they would refuse.

### Bivariate determinants of vaccine acceptance

All sociodemographic characteristics examined were significantly associated with acceptance status (Table 1). Acceptance declined steadily with age, from 88.4% among 18–24-year-olds to 53.1% among those aged 45 and above (p < 0.001). A clear educational gradient was evident, though non-linear: acceptance was lowest among respondents with no or only primary education (28.6%), rose sharply among those with Junior High School (84.7%) and Secondary education (90.5%), and was comparatively lower again among tertiary-educated respondents (65.9%) (p < 0.001). Occupation showed a similarly strong pattern, with students (94.3%) and private-sector workers (93.6%) showing the highest acceptance and the unemployed/retired group the lowest (28.6%) (p < 0.001). Acceptance also differed by peri-urban area, ranging from 55.1% in Adiembra to 95.6% in Kojokrom (p < 0.001).

All HBM construct items were significantly associated with acceptance status (Table 2). The steepest gradient was observed for perceived vaccine safety: acceptance ranged from 95.5% among those who considered the vaccine “very safe” to 14.7% among those who considered it “not safe,” with refusal rising correspondingly from 2.4% to 58.8% (p < 0.001). A similarly steep gradient was observed for perceived benefits (89.5% acceptance among those believing the vaccine effective, versus 48.6% among those who did not, p < 0.001). Perceived susceptibility and perceived severity showed non-monotonic patterns: respondents at the extremes (either clearly at risk/concerned, or clearly not) showed higher acceptance than those in the intermediate “not sure”/“somewhat concerned” categories, suggesting that ambivalence itself, and not merely low risk perception, is associated with reduced acceptance. Knowledge was also associated with acceptance status (p < 0.001): acceptance was 87.6% among respondents with good knowledge, 76.2% and 73.9% among those with moderate and poor knowledge, and 42.4% among those who had not heard of mpox. The association persisted when the analysis was restricted to the 495 respondents who had heard of mpox (p = 0.007).

### Multivariable determinants of vaccine acceptance

Multinomial logistic regression was used to identify independent determinants of hesitancy and refusal, relative to acceptance, adjusting simultaneously for sociodemographic characteristics, HBM constructs, and peri-urban area (Table 3; full results in S1 Table). The model fit the data well (McFadden pseudo-R² = 0.51; likelihood ratio test p < 0.001). Both perceived vaccine safety (likelihood ratio ² = 48.0, df = 6, p < 0.001) and peri-urban area ( ² = 27.2, df = 10, p = 0.002) contributed significantly to model fit, whereas adding knowledge did not ( ² = 3.4, df = 6, p = 0.76). There was no evidence of quasi-separation (maximum standard error 1.71), and all variance inflation factors were below 7.

**Table 3.**
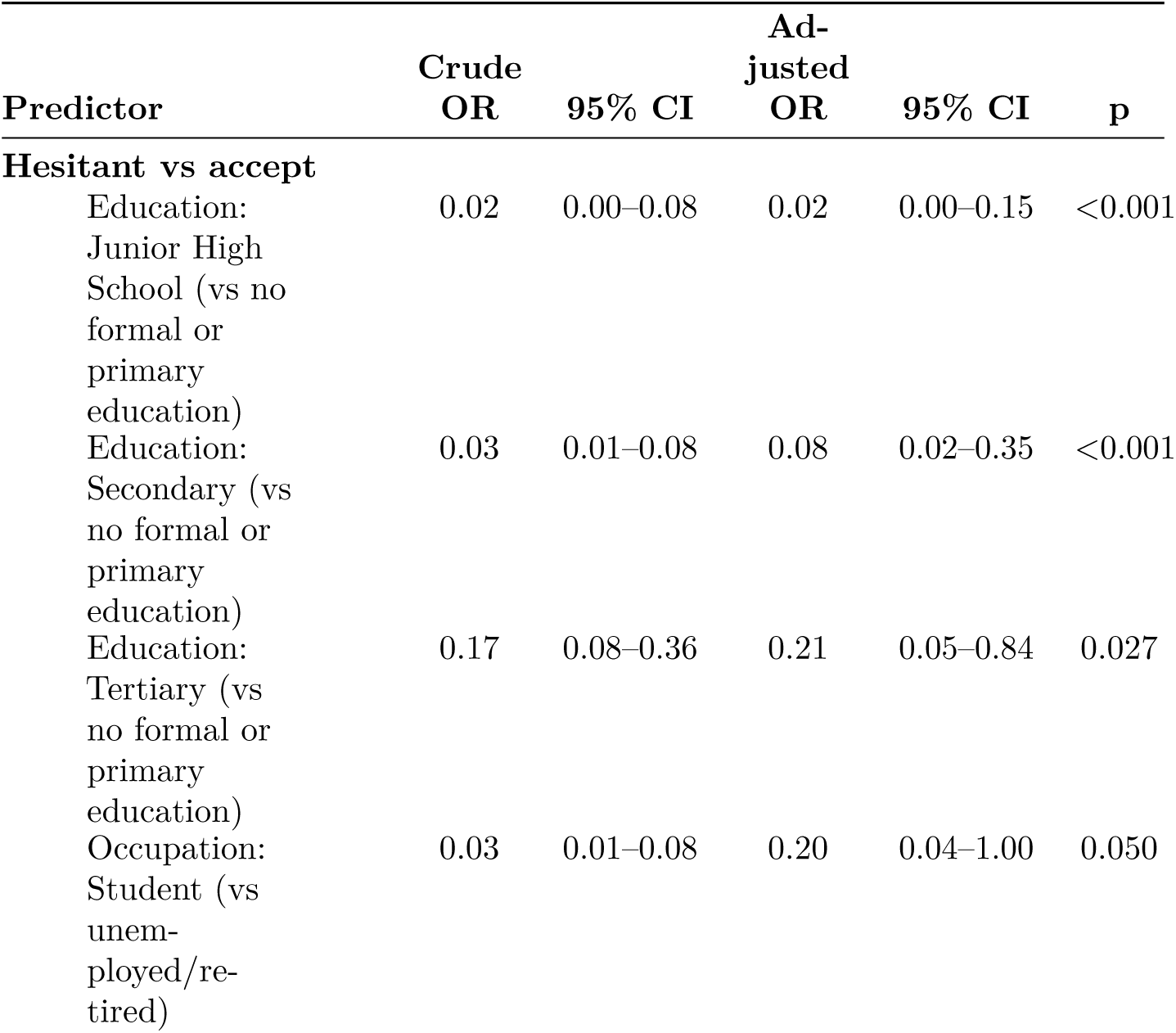
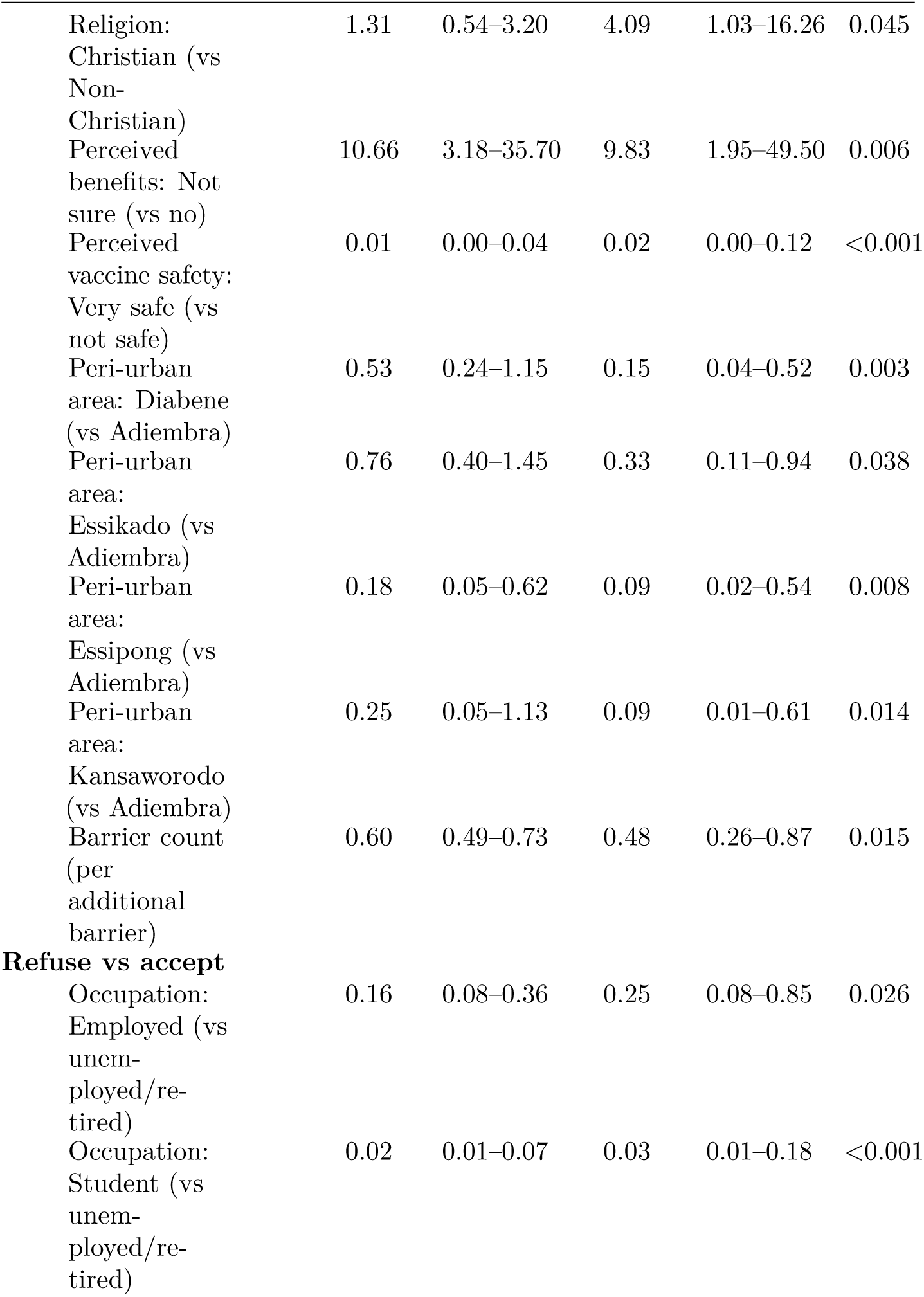
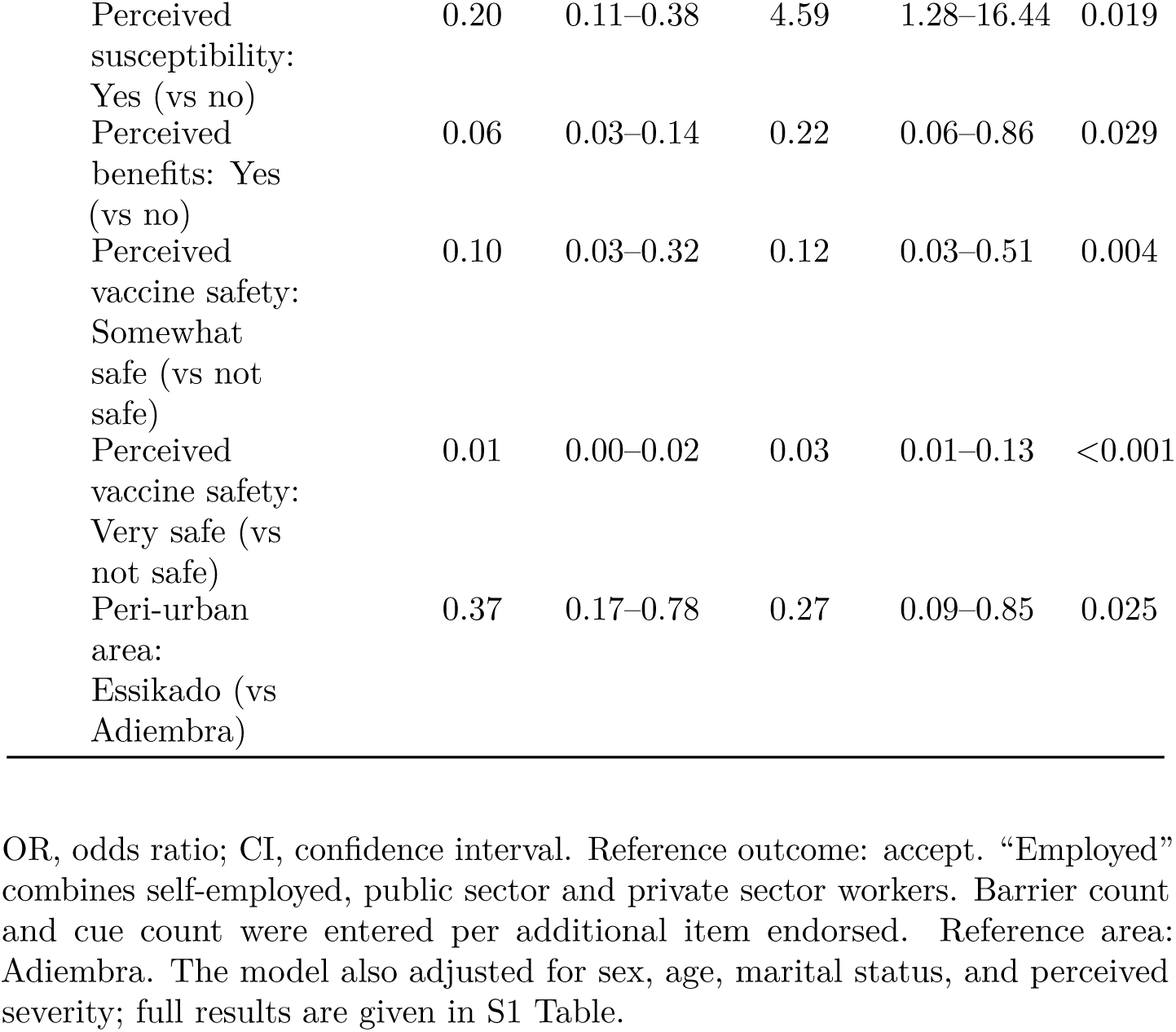
Crude and adjusted odds ratios from multinomial logistic regression for the statistically significant predictors (p < 0.05).

| **Predictor** | **Crude OR** | **95% CI** | **Ad-justed OR** | **95% CI** | **p** |
|---|---|---|---|---|---|
| **Hesitant vs accept** |  |  |  |  |  |
| Education: Junior High School (vs no formal or primary education) | 0.02 | 0.00–0.08 | 0.02 | 0.00–0.15 | <0.001 |
| Education: Secondary (vs no formal or primary education) | 0.03 | 0.01–0.08 | 0.08 | 0.02–0.35 | <0.001 |
| Education: Tertiary (vs no formal or primary education) | 0.17 | 0.08–0.36 | 0.21 | 0.05–0.84 | 0.027 |
| Occupation: Student (vs unem-ployed/re-tired) | 0.03 | 0.01–0.08 | 0.20 | 0.04–1.00 | 0.050 |
| Religion: Christian (vs Non-Christian) | 1.31 | 0.54–3.20 | 4.09 | 1.03–16.26 | 0.045 |
| Perceived benefits: Not sure (vs no) | 10.66 | 3.18–35.70 | 9.83 | 1.95–49.50 | 0.006 |
| Perceived vaccine safety: Very safe (vs not safe) | 0.01 | 0.00–0.04 | 0.02 | 0.00–0.12 | <0.001 |
| Peri-urban area: Diabene (vs Adiembra) | 0.53 | 0.24–1.15 | 0.15 | 0.04–0.52 | 0.003 |
| Peri-urban area: Essikado (vs Adiembra) | 0.76 | 0.40–1.45 | 0.33 | 0.11–0.94 | 0.038 |
| Peri-urban area: Essipong (vs Adiembra) | 0.18 | 0.05–0.62 | 0.09 | 0.02–0.54 | 0.008 |
| Peri-urban area: Kansaworodo (vs Adiembra) | 0.25 | 0.05–1.13 | 0.09 | 0.01–0.61 | 0.014 |
| Barrier count (per additional barrier) | 0.60 | 0.49–0.73 | 0.48 | 0.26–0.87 | 0.015 |
| **Refuse vs accept** |  |  |  |  |  |
| Occupation: Employed (vs unem-ployed/re-tired) | 0.16 | 0.08–0.36 | 0.25 | 0.08–0.85 | 0.026 |
| Occupation: Student (vs unem-ployed/re-tired) | 0.02 | 0.01–0.07 | 0.03 | 0.01–0.18 | <0.001 |
| Perceived susceptibility: Yes (vs no) | 0.20 | 0.11–0.38 | 4.59 | 1.28–16.44 | 0.019 |
| Perceived benefits: Yes (vs no) | 0.06 | 0.03–0.14 | 0.22 | 0.06–0.86 | 0.029 |
| Perceived vaccine safety: Somewhat safe (vs not safe) | 0.10 | 0.03–0.32 | 0.12 | 0.03–0.51 | 0.004 |
| Perceived vaccine safety: Very safe (vs not safe) | 0.01 | 0.00–0.02 | 0.03 | 0.01–0.13 | <0.001 |
| Peri-urban area: Essikado (vs Adiembra) | 0.37 | 0.17–0.78 | 0.27 | 0.09–0.85 | 0.025 |

OR, odds ratio; CI, confidence interval. Reference outcome: accept. “Employed” combines self-employed, public sector and private sector workers. Barrier count and cue count were entered per additional item endorsed. Reference area: Adiembra. The model also adjusted for sex, age, marital status, and perceived severity; full results are given in S1 Table.

#### Determinants of hesitancy (vs. acceptance)

Perceived vaccine safety was the strongest determinant. Compared with respondents who considered the vaccine not safe, those who considered it very safe had 98% lower adjusted odds of being hesitant rather than accepting (aOR 0.02, 95% CI 0.00–0.12, p < 0.001). Respondents who were unsure of the vaccine’s effectiveness had higher adjusted odds of hesitancy than those who believed it ineffective (aOR 9.83, 95% CI 1.95–49.50, p = 0.006). Compared with respondents with no formal or primary education, those with Junior High School (aOR 0.02, p < 0.001), secondary (aOR 0.08, p < 0.001), and tertiary (aOR 0.21, p = 0.027) education had lower odds of hesitancy; the association for students relative to unemployed or retired respondents was borderline (aOR 0.20, 95% CI 0.04–1.00, p = 0.050). Christian respondents had higher odds of hesitancy than non-Christian respondents (aOR 4.09, 95% CI 1.03–16.26, p = 0.045), and each additional barrier endorsed was associated with lower odds of hesitancy (aOR 0.48, 95% CI 0.26–0.87, p = 0.015). Compared with Adiembra, the adjusted odds of hesitancy were lower in Diabene (aOR 0.15, p = 0.003), Essikado (aOR 0.33, p = 0.038), Essipong (aOR 0.09, p = 0.008), and Kansaworodo (aOR 0.09, p = 0.014).

#### Determinants of refusal (vs. acceptance)

Perceived safety was again the dominant determinant. Relative to respondents who considered the vaccine not safe, the adjusted odds of refusal were lower among those who considered it very safe (aOR 0.03, 95% CI 0.01–0.13, p < 0.001) or somewhat safe (aOR 0.12, 95% CI 0.03–0.51, p = 0.004). Belief in vaccine effectiveness remained independently protective against refusal (aOR 0.22, 95% CI 0.06–0.86, p = 0.029), as did being a student (aOR 0.03, p < 0.001) or employed (aOR 0.25, p = 0.026) rather than unemployed or retired. Residents of Essikado had lower odds of refusal than residents of Adiembra (aOR 0.27, 95% CI 0.09–0.85, p = 0.025). Unexpectedly, respondents who considered themselves at risk of mpox had higher adjusted odds of refusal than those who did not (aOR 4.59, 95% CI 1.28–16.44, p = 0.019), the reverse of the crude association (crude OR 0.20).

Age, sex, and marital status, which showed significant crude associations with acceptance status, were not independently associated with it after adjustment, suggesting that their bivariate associations largely reflected their correlation with education and occupation. Perceived severity and barrier count were associated with refusal in a model without perceived safety (S3 Table) but not once safety was included, and the cue-to-action count was associated with refusal in a model without peri-urban area (S2 Table) but not once area was included.

## Discussion

### Summary of key findings

This study found that three-quarters of peri-urban residents in the Sekondi-Takoradi Metropolis (75.7%) were willing to accept the mpox vaccine, with 12.9% hesitant and 11.4% refusing. Beliefs about the vaccine itself, namely its safety and its effectiveness, were the strongest determinants of acceptance: perceived safety was independently associated with both hesitancy and refusal, uncertainty about effectiveness with hesitancy, and belief in effectiveness with lower odds of refusal. Education and occupation were also independently associated with acceptance status, and acceptance differed substantially between peri-urban areas. Knowledge of mpox transmission was associated with acceptance in bivariate analysis but not after adjustment for vaccine beliefs and sociodemographic characteristics.

### Vaccine acceptance in context

The acceptance rate observed here is notably higher than in earlier Ghanaian mpox studies. Ghazy et al. [17], surveying a national online sample in late 2022, found that only 46.1% of Ghanaians intended to accept the mpox vaccine, and Braimah et al. [18], using cross-sectional data from four major Ghanaian cities, reported an even lower acceptance rate of approximately 32%. A plausible explanation lies in timing and context. Data for the present study were collected in July–August 2025, during an escalating outbreak in which the Western Region had become the epicentre of Ghana’s mpox cases, and before the sub-national vaccination campaign was launched in October 2025 [6]. Respondents were therefore living through a visible, community-level outbreak and were exposed to outbreak-related health messaging, including risk communication and health education, rather than responding to a hypothetical or distant threat. Direct proximity to a live outbreak plausibly heightened perceived susceptibility and severity in ways that earlier, pre-outbreak surveys could not capture, consistent with the Health Belief Model’s premise that perceived threat is a precondition for protective action. This pattern also echoes Elsayed et al. [38], who found that recent, lived outbreak exposure shaped vaccine-relevant beliefs in Dar es Salaam more than background knowledge did. Because the data predate the campaign, these findings also serve as a pre-campaign baseline against which uptake during and after the campaign can be compared.

### Health Belief Model constructs and acceptance

Perceived vaccine safety and perceived benefits were the dominant constructs in this study. Respondents who regarded the vaccine as very safe had 98% and 97% lower adjusted odds of hesitancy and refusal, respectively, than those who regarded it as unsafe, and fear of side effects was the most frequently endorsed concern (79.1%). The importance of perceived benefits is consistent with the broader HBM-vaccine literature. Riad et al. [28], studying mpox vaccine hesitancy among Belarusian healthcare professionals, identified perceived benefits as the construct most strongly correlated with willingness to vaccinate, and Mohamed et al. [27], applying the HBM to mpox vaccine acceptance among Gulf healthcare workers, found perceived benefits and self-efficacy to be the strongest predictors. Al-Metwali et al. [39] reached the same conclusion for COVID-19 vaccine acceptance in Iraq, where perceived benefit was among the most robust correlates identified. In the present study, the protective association between belief in vaccine effectiveness and refusal was attenuated once perceived safety was taken into account but remained significant (aOR 0.22, 95% CI 0.06–0.86). The two beliefs therefore overlap substantially, since most respondents who considered the vaccine very safe also considered it effective, yet each contributes independently.

Perceived susceptibility behaved differently. In bivariate analysis, respondents who considered themselves at risk were more likely to accept the vaccine, but after adjustment they had higher odds of refusal than those who did not consider themselves at risk (aOR 4.59, 95% CI 1.28–16.44). This reversal, together with the wide confidence interval, suggests that once beliefs about vaccine safety and effectiveness are held constant, perceived risk adds little independent motivation to be vaccinated, and the finding should be interpreted cautiously. A related pattern was reported by Wong et al. [40], who, in a population-based HBM study of COVID-19 vaccine acceptance in Hong Kong, found that perceived susceptibility carried no significant association with acceptance once other constructs were considered; the authors attributed this to governmental recommendation and perceived benefit overriding personal risk calculus. The outbreak’s visibility likely elevated baseline risk perception across nearly the entire sample (74.0% considered themselves at risk), leaving relatively little variance for susceptibility to explain once vaccine beliefs were accounted for.

The non-monotonic pattern observed for both susceptibility and severity, in which the intermediate “not sure” and “somewhat concerned” categories showed lower acceptance than either extreme, is a less commonly reported finding in this literature, though it aligns conceptually with treating hesitancy as a distinct psychological state rather than as a midpoint on a single continuum from acceptance to refusal, consistent with the working group definition adopted here and with this study’s decision to model acceptance as three categories rather than collapsing to binary. It suggests that ambivalence about personal risk, not simply low perceived risk, is itself an obstacle to firm acceptance, a nuance that binary-outcome studies in this literature would not be able to detect.

The barrier and cue-to-action counts require careful interpretation. The barrier question was worded for respondents who were unwilling or unsure but was answered by all respondents, and accepters endorsed more concerns on average than hesitant or refusing respondents (mean 2.6, 1.7, and 2.0 concerns, respectively). After adjustment, the barrier count was not associated with refusal (aOR 1.28, 95% CI 0.74–2.21), while a higher count was associated with lower odds of hesitancy, which likely reflects accepters listing hypothetical concerns rather than barriers that deterred them. The cue-to-action count was not independently associated with either outcome once peri-urban area was taken into account. Descriptively, refusers named the fewest factors that would increase their willingness to vaccinate (mean 1.7, compared with 2.7 among accepters) and most often cited mistrust in vaccines (76%), consistent with firmer opposition that is less open to persuasion, whereas accepters and hesitant respondents pointed to concrete needs, namely more information from health authorities, assurance of safety and effectiveness, and visible peer uptake. These factors are better read as a guide to where targeted intervention could still move undecided individuals than as determinants in their own right, and the results caution against interpreting count-based HBM measures without multivariable adjustment.

Knowledge of mpox transmission was associated with acceptance in bivariate analysis: acceptance was 87.6% among respondents with good knowledge but 42.4% among those who had not heard of mpox. However, knowledge was not independently associated with acceptance once sociodemographic characteristics, vaccine beliefs, and area were taken into account (likelihood ratio test p = 0.76). This aligns with Elsayed et al. [38], who, examining knowledge, attitudes, and practices toward viral disease outbreaks including mpox in Tanzania, concluded that vaccine hesitancy was driven primarily by concerns about efficacy and fear of side effects rather than by overall knowledge deficits. Taken together, these findings suggest that awareness of the disease is a necessary foundation, since respondents who had never heard of mpox were the least willing to be vaccinated, but that transmission-route education alone is unlikely to be sufficient; beliefs about the vaccine itself appear to be the more direct intervention target.

### Sociodemographic determinants

Education showed a distinctive non-linear relationship with acceptance: respondents with Junior High School or Secondary education showed markedly higher acceptance than either those with no/primary education or, notably, those with tertiary education. This is a departure from the assumption that acceptance rises monotonically with education, but it is not without precedent: higher educational attainment can be associated with greater exposure to competing or conflicting information sources and more independent critical evaluation of health claims, which does not uniformly translate into higher acceptance. The occupational gradient was more conventional: students and the formally employed showed substantially higher acceptance than the unemployed or retired, consistent with Bushi et al.’s [21] systematic review of malaria vaccine acceptance determinants across sub-Saharan Africa, which similarly identified employment status as a significant correlate of vaccine acceptance, plausibly reflecting differential access to health information, healthcare services, and institutional trust associated with formal engagement in education or work.

Christian respondents had higher adjusted odds of hesitancy than non-Christian respondents (aOR 4.09, 95% CI 1.03–16.26), with no corresponding difference in outright refusal. Descriptively, Muslim respondents were more polarised (a higher refusal rate but a lower hesitancy rate) than Christian respondents, who more often occupied the undecided middle category. Given that the non-Christian reference group was small (n = 69, predominantly Muslim) and the confidence interval wide, this pattern is better treated as a hypothesis for qualitative investigation into denomination-specific decision-making, of the kind explored by Aberese-Ako et al. [24] in their ethnographic study of community engagement and COVID-19 vaccine acceptance in Ghana, than as a settled conclusion.

Acceptance also differed substantially between peri-urban areas, from 55.1% in Adiembra to 95.6% in Kojokrom. Part of this difference reflects the composition of the samples: 61% of Kojokrom respondents were students, compared with 13–25% in the other areas. After adjustment for individual characteristics and vaccine beliefs, residents of Diabene, Essikado, Essipong, and Kansaworodo remained less likely than residents of Adiembra to be hesitant, and Essikado residents were also less likely to refuse. These residual differences may reflect local variation in outbreak exposure, health messaging, or community leadership, and suggest that vaccination outreach should be tailored to individual communities rather than applied uniformly across the metropolis.

Notably, age, sex, and marital status all showed strong, statistically significant associations with acceptance status in bivariate analysis, but none remained significant after adjustment for education, occupation, religion, and the HBM constructs. This indicates that their apparent influence operated indirectly, through their correlation with education and occupation, rather than as independent determinants, a distinction only visible because both crude and adjusted estimates were reported, and a reminder that single-variable descriptive comparisons common in much of the vaccine-acceptance literature can overstate the independent role of basic demographic characteristics.

### Strengths and limitations

This study’s principal strength is its timing and specificity: it captures vaccine-related beliefs from a defined peri-urban population during an active, geographically concentrated outbreak and before vaccination began, using a theoretically grounded instrument and a three-category outcome that preserves the distinction between hesitancy and refusal, a distinction most binary-outcome studies in this literature cannot make. Several limitations should be acknowledged. First, the peri-urban areas were purposively selected and respondents were recruited from among available adults rather than by random selection; the sample over-represents women (70.0%) and students (33.1%), the level of acceptance may therefore not reflect the wider peri-urban population, and the sample size calculation did not account for a design effect. Second, respondents were clustered within six areas; area was included as a fixed effect, but with so few areas, between-area variation could not be modelled as a random effect. Third, most HBM constructs were measured with single items, self-efficacy was not measured, and the barrier question was answered by all respondents although it was worded for those who were unwilling or unsure. Fourth, the cross-sectional design precludes causal inference; the associations identified here describe correlates of stated willingness at a single point in time rather than confirmed drivers of vaccination behaviour. Fifth, self-reported willingness may be subject to social desirability bias, particularly during an active outbreak with prominent public health messaging. Finally, generalisation to urban-core or rural populations elsewhere in the Western Region, or to Ghana more broadly, should be made cautiously.

## Conclusions

This study found that three-quarters of peri-urban residents of the Sekondi-Takoradi Metropolis were willing to accept the mpox vaccine during an active regional outbreak, before the vaccination campaign began. Beliefs about the vaccine itself were the strongest determinants: perceived safety was independently associated with both hesitancy and refusal, and beliefs about its effectiveness with both outcomes. Education, occupation, and peri-urban area also independently shaped acceptance status, whereas knowledge of transmission, although associated with acceptance in bivariate analysis, was not independently associated with it once vaccine beliefs were considered. By modelling acceptance as three categories rather than collapsing it to a binary outcome, this study showed that hesitancy and outright refusal are not driven by identical factors: hesitancy was most closely tied to uncertainty about vaccine benefit, safety concerns, and lower education, while refusal was shaped by safety concerns, disbelief in vaccine effectiveness, and employment status.

These findings suggest that mpox vaccination strategies in similar peri-urban West African settings should prioritise clear, credible communication about vaccine safety and effectiveness, building on basic disease awareness rather than relying on it, and should be deliberately targeted toward unemployed and less-educated residents, who showed disproportionately lower acceptance independent of their underlying beliefs, as well as toward areas with lower acceptance. Because data collection coincided with a live, geographically concentrated outbreak and preceded the vaccination campaign, these results offer a rare within-outbreak, pre-campaign baseline of how vaccine-related beliefs behave under real, rather than hypothetical, threat conditions. This evidence can inform the vaccination response in the Western Region and future outbreak preparedness planning across similar communities in Ghana and the wider sub-region.

## Data Availability

The minimal de-identified dataset and accompanying analysis code underlying this study are available at Zenodo via https://doi.org/10.5281/zenodo.24000845

## Acknowledgments

We acknowledge the Western Regional Health Directorate and the Sekondi-Takoradi Metropolitan Health Directorate of the Ghana Health Service, and all data collection officers engaged to undertake this work.

## Supporting information

**S1 Table. Full multinomial logistic regression results.** Coefficients, standard errors, adjusted and crude odds ratios with 95% confidence intervals, and p-values for all predictors in the main model.

**S2 Table. Multinomial logistic regression model without peri-urban area.** Crude and adjusted odds ratios from the model that excluded peri-urban area, for comparison with the main model.

**S3 Table. Multinomial logistic regression model without perceived vaccine safety.** Crude and adjusted odds ratios from the model that excluded perceived vaccine safety, for comparison with the main model.

**S1 Code. Python analysis code.** Code used for data preparation, descriptive and bivariate analysis, and multinomial logistic regression.

## Notes

### Competing Interest Statement

The authors have declared no competing interest.

### Author Declarations

Ethical approval for this study was obtained from the University Research Ethics Committee of Takoradi Technical University, Takoradi, Ghana (reference number TTU/UREC/26/097, approved 17 June 2025). Permission to conduct the study was also obtained from the Sekondi-Takoradi Metropolitan Health Directorate, Ghana Health Service. Verbal informed consent was obtained from all participants and recorded electronically before data collection.

## References

1. Cordeiro R, Francisco R, Pelerito A, Lopes de Carvalho I, Núncio MS. Mpox surveillance and laboratory response in Portugal: Lessons learned from three outbreak waves (2022–2025). Infect Dis Rep. 2025;17(4):86. doi: 10.3390/idr17040086

2. Satapathy P, Yoosuf BT, Gaidhane AM, Vadia N, Menon SV, Chennakesavulu K, et al. Mpox: Transmission dynamics, treatment, and innovations. Ther Adv Infect Dis. 2025;12:20499361251357521. doi: 10.1177/20499361251357521

3. Araf Y, Nipa JF, Naher S, Maliha ST, Rahman H, Arafat KI, et al. Insights into the transmission, host range, genomics, vaccination, and current epidemiology of the monkeypox virus. Vet Med Int. 2024;2024:8839830. doi: 10.1155/2024/8839830

4. World Health Organization. Multi-country outbreak of mpox: External situation report no. 60, 23 November 2025. Geneva: World Health Organization; 2025 [cited 2026 Nov 2]. Available from: https://cdn.who.int/media/docs/default-source/documents/emergencies/multi-country-outbreak-of-mpox--external-situation-report--60.pdf

5. Aabalekuu S. Mpox outbreak in Ghana: Epidemiology, response, and lessons learned. Frontline Professionals Journal. 2026;3(2):50–58. doi: 10.60787/fpj.vol3no2.112

6. Ghana Health Service. Ghana Health Service launches mpox vaccination campaign in Takoradi [Internet]. Accra: Ghana Health Service; 2025 Oct 5 [cited 2026 Nov 4]. Available from: https://ghs.gov.gh/news-and-events/ghana-health-service-launches-mpox-vaccination-campaign-in-takoradi

7. Danladi NP, Agboola P, Olaniyi P, Eze S, Oladapo O, Obiwulu D, et al. Challenges in global distribution and equitable access to monkeypox vaccines. Viruses. 2024;16(12):1815. doi: 10.3390/v16121815

8. Chandipwisa C, Shimilimo A, Uwishema O, Onyeaka H. The current landscape and future prospects of vaccine manufacturing in Africa: Challenges, innovations, and opportunities: A narrative review. Health Sci Rep. 2026;9(3):e71926. doi: 10.1002/hsr2.71926

9. Abiye AA, Endale S, Negash Z, Hussen SU, Gebregeorgise DT, Tamiru MT, et al. COVID-19 vaccine hesitancy, self-reported adverse events, and determinants among Ethiopian healthcare professionals during COVID-19 pandemic. Public Health Chall. 2025;4(3):e70078. doi: 10.1002/puh2.70078

10. Riad A, Drobov A, Rozmarinová J, Drapáčová P, Klugarová J, Dušek L, et al. Monkeypox knowledge and vaccine hesitancy of Czech healthcare workers: A Health Belief Model (HBM)-based study. Vaccines (Basel). 2022;10(12):2022. doi: 10.3390/vaccines10122022

11. Hong J, Pan B, Jiang H, Zhang Q, Xu X, Jiang H, et al. The willingness of Chinese healthcare workers to receive monkeypox vaccine and its independent predictors: A cross-sectional survey. J Med Virol. 2022;95(1):e28294. doi: 10.1002/jmv.28294

12. Dukers-Muijrers NHTM, Evers Y, Widdershoven V, Davidovich U, Adam PCG, Op de Coul ELM, et al. Mpox vaccination willingness, determinants, and communication needs in gay, bisexual, and other men who have sex with men, in the context of limited vaccine availability in the Netherlands (Dutch Mpox-survey). Front Public Health. 2023;10:1058807. doi: 10.3389/fpubh.2022.1058807

13. Li Y, Peng X, Fu L, Wang B, Sun Y, Chen Y, et al. Monkeypox awareness and low vaccination hesitancy among men who have sex with men in China. J Med Virol. 2023;95(2):e28567. doi: 10.1002/jmv.28567

14. Mohamed MG, Rasheed HM, Bhagavathy MG, Wishah MA, Ghosh DP, Islam MR, et al. Decoding vaccine hesitancy in mpox across multiple Middle Eastern countries: Public knowledge, attitudes, worry, conspiracy beliefs, and acceptance. Health Sci Rep. 2026;9(5):e72444. doi: 10.1002/hsr2.72444

15. Caycho-Rodríguez T, Valencia PD, Ventura-León J, Carbajal-León C, Vilca LW, Gallegos M, et al. Intention to receive the monkeypox vaccine and its psychological and sociodemographic predictors: A cross-sectional survey in the general population of Peru. Psicol Reflex Crit. 2023;36(1):39. doi: 10.1186/s41155-023-00281-z

16. Peptan C, Mărcău FC, Alecsoiu OR, Panagoret DM, Cojoaca ME, Musetescu AM, et al. The evolution of monkeypox vaccination acceptance in Romania: A comparative analysis (2022–2025), psychosocial perceptions, and the impact of anti-vaccination rhetoric on societal security. Behav Sci (Basel). 2025;15(9):1175. doi: 10.3390/bs15091175

17. Ghazy RM, Yazbek S, Gebreal A, Hussein M, Addai SA, Mensah E, et al. Monkeypox vaccine acceptance among Ghanaians: A call for action. Vaccines (Basel). 2023;11(2):240. doi: 10.3390/vaccines11020240

18. Braimah JA, Achore M, Dery F, Ayanore MA, Bisung E, Kuuire V. Do self-rated health and previous vaccine uptake influence the willingness to accept MPOX vaccine during a public health emergency of concern? A cross-sectional study. PLOS Glob Public Health. 2024;4(8):e0003564. doi: 10.1371/journal.pgph.0003564

19. Ghazy RM, Okeh DU, Sallam M, Hussein M, Ismail HM, Yazbek S, et al. Psychological antecedents of healthcare workers towards monkeypox vaccination in Nigeria. Vaccines (Basel). 2022;10(12):2151. doi: 10.3390/vaccines10122151

20. Iliyasu Z, Abdullahi HM, Umar AA, Dahiru T, Tsiga-Ahmed FI, Amole TG, et al. Public perception and correlates of Mpox vaccine acceptability in northern Nigeria: A mixed methods study. Pan Afr Med J. 2025;50(Suppl 1):4. doi: 10.11604/pamj.supp.2025.50.1.44316

21. Bushi G, Khatib MN, S RJ, Kaur I, Sharma A, Iqbal S, et al. Determinants of malaria vaccine acceptance: A systematic review and meta-analysis of awareness, acceptance, hesitancy, and willingness to pay. Immun Inflamm Dis. 2025;13(5):e70205. doi: 10.1002/iid3.70205

22. Aram SA, Hagan JE, Mansoh GK, Saalidong BM, Lartey PO, Ahinkorah BO, et al. Association between health and safety perceptions of COVID-19 vaccine and its uptake in Ghana. Public Health Chall. 2022;1(4):e20. doi: 10.1002/puh2.20

23. Tabong PT, Opoku Mensah K, Asampong E. Preparation for COVID-19 vaccines rollout: Interventions to increase trust, acceptability, and uptake in West African countries. Int J Health Plann Manage. 2022;37(3):1221–1228. doi: 10.1002/hpm.3426

24. Aberese-Ako M, Ebelin W, Doegah PT, Kuatewo M, Kpodo L, Kpordorlor AG, et al. Promoting COVID-19 vaccine acceptance through community engagement: An ethnographic study in Ghana. Adv Public Health. 2023;2023:3626862. doi: 10.1155/2023/3626862

25. Aninagyei E, Ayivor-Djanie R, Boatey CA, Mateen AA, Awuku SA, Boakye AA. High SARS-CoV-2 antibody seroprevalence amid COVID-19 vaccine hesitancy among unvaccinated adults in Ghana. J Med Virol. 2026;98(5):e70962. doi: 10.1002/jmv.70962

26. Pan WK. The application of behavioral change theories in addressing vaccine hesitancy: A literature review. Public Health Nurs. 2024;41(2):318–324. doi: 10.1111/phn.13280

27. Mohamed MG, Dabou EAA, Ibrahim FM, Abdsalam SAE, Kunjavara J, Mannethodi K, et al. Assessment of healthcare workers’ knowledge and attitudes toward mpox and acceptance of its vaccine: A health belief model-based analysis in the Gulf region. Osong Public Health Res Perspect. 2025;16(4):357–372. doi: 10.24171/j.phrp.2025.0113

28. Riad A, Rybakova N, Dubatouka N, Zankevich I, Klugar M, Koščík M, et al. Belarusian healthcare professionals’ views on monkeypox and vaccine hesitancy. Vaccines (Basel). 2023;11(8):1368. doi: 10.3390/vaccines11081368

29. Tweneboah-Koduah EY. Social marketing: Using the health belief model to understand breast cancer protective behaviours among women. Int J Nonprofit Volunt Sect Mark. 2018;23(2):e1613. doi: 10.1002/nvsm.1613

30. Kyei-Gyamfi S, Kyei-Gyamfi A, Owusu K, Awuviri I, Charway GA, Kwatsenu J, et al. Condom use among internal migrant girls in Ghana: Exploring the barriers, enablers, and willingness to use among sexually active Kayayei. Reprod Female Child Health. 2025;4(4):e70041. doi: 10.1002/rfc2.70041

31. Bawuah A, Wang VQ, Adeleye K, Ameyaw EK, Yaya S. Mass media exposure and cervical cancer screening in rural sub-Saharan Africa: A multi-country cross-sectional analysis. Health Sci Rep. 2025;8(4):e70663. doi: 10.1002/hsr2.70663

32. Shen Z, Li Q, Wu J, Zhu D, Bai J, Ren R, et al. Dynamic evolution of COVID-19 vaccine hesitancy over 2021–2023 among Chinese population: Repeated nationwide cross-sectional study. J Med Virol. 2024;96(7):e29800. doi: 10.1002/jmv.29800

33. Owens C, Hubach RD. Rural-urban differences in monkeypox behaviors and attitudes among men who have sex with men in the United States. J Rural Health. 2022;39(2):508–515. doi: 10.1111/jrh.12726

34. Vasudevan L, Wang Y, Ostermann J, Yelverton V, Yang J, Fish LJ, et al. Rural-urban disparities in human papillomavirus vaccination: Findings from a cross-sectional survey of 13 southern US states, December 2019–January 2020. J Rural Health. 2025;41(2):e12913. doi: 10.1111/jrh.12913

35. Cissé K, Zoungrana EI, Zouré AA, Sagna T, Tao G, Zida S, et al. Determinants of COVID-19 vaccination status and vaccine hesitancy in the Centre-Est Region of Burkina Faso: A population-based cross-sectional study. Adv Public Health. 2025;2025:8152436. doi: 10.1155/adph/8152436

36. Oladapo RK, Obidiro OP, Oyetola AB, Olajide A, Oladapo MO, Lawrence US, et al. COVID-19 vaccination in low and middle-income countries: Creating a sustainable roadmap for promoting public health intervention. Int J Health Plann Manage. 2024;39(5):1612–1625. doi: 10.1002/hpm.3834

37. Ghana Statistical Service. Ghana 2021 Population and Housing Census. General Report Volume 3A: Population of Regions and Districts. Accra: Ghana Statistical Service; 2021. Available from: https://statsghana.gov.gh/gssmain/file-Upload/pressrelease/2021%20PHC%20General%20Report%20Vol%203A_Population%20of%20Regions%20and%20Districts_181121.pdf

38. Elsayed MEF, Magandi EJ, Rugarabamu S. Community awareness, knowledge, attitudes, and perceptions toward viral disease outbreaks in Dar es Salaam, Tanzania. Public Health Chall. 2026;5(2):e70232. doi: 10.1002/puh2.70232

39. Al-Metwali BZ, Al-Jumaili AA, Al-Alag ZA, Sorofman B. Exploring the acceptance of COVID-19 vaccine among healthcare workers and general population using health belief model. J Eval Clin Pract. 2021;27(5):1112–1122. doi: 10.1111/jep.13581

40. Wong MCS, Wong ELY, Huang J, Cheung AWL, Law K, Chong MKC, et al. Acceptance of the COVID-19 vaccine based on the health belief model: A population-based survey in Hong Kong. Vaccine. 2021;39(7):1148–1156. doi: 10.1016/j.vaccine.2020.12.083

